# Unweighted versus weighted regression methods may be sufficient to analyze complex survey data in the Canadian Longitudinal Study on Aging

**DOI:** 10.1101/2022.08.31.22279464

**Authors:** Mark Oremus, Colleen Maxwell, Suzanne L. Tyas, Lauren E. Griffith, Edwin R. van den Heuvel

## Abstract

**Introduction:** Complex surveys use stratified or cluster sampling to recruit participants. Researchers analyzing these surveys often wish to make inferences about the source populations from which the participants are drawn. In such cases, methodologists recommend employing sample weights in regression analyses; however, the utilization of weights in studies of associations are not without dispute.

**Materials and methods:** To help guide analyses of complex surveys, we utilized baseline data from the Comprehensive Cohort of the Canadian Longitudinal Study on Aging (CLSA) (n = 30,097) and compared unweighted and weighted regression analyses of the association between social support availability (SSA) and cognitive function. We also conducted simulation studies to validate our findings in the CLSA.

**Results:** The regression coefficients 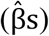 for SSA were similar across the unweighted and weighted regression models; the standard errors of the 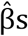 were lower in the unweighted models. The simulation studies mimicking CLSA confirmed these findings. Overall, our findings demonstrated a small advantage for the unweighted analysis of CLSA data due to the smaller standard errors.

**Discussion:** Although we cannot guarantee that this would be the case for all association analyses with CLSA data, the current study showed the use of analytical weights was not necessary for our associations of interest.

## Introduction

In surveys involving stratified or cluster sampling, individuals in the source populations often have unequal probabilities of being selected to participate. Researchers who work with data from these so-called ‘complex’ surveys must choose whether to incorporate sample weights into their analyses. Each survey participant’s numeric sample weight is an estimate of the number of people in the source population who are represented by that participant. Prevalence estimates, means, and regression coefficients obtained from weighted analyses apply to all of the individuals in a given source population, not just to survey participants [1,2]. However, researchers sometimes ignore sample weights, even when they wish to use survey data to draw inferences about a source population, and treat complex survey data as simple random samples with no error or bias. This is equivalent to setting each participant’s weight to one. Analysts sometimes prefer to ignore weights when the calculation of the weights is opaque or the intricacies of the sampling process are unclear [3,4].

Recommendations to use weights when analyzing complex surveys have existed for over three decades [2], and the debate over using weights in regression analyses of data from such surveys has existed for almost as long, with equivocal conclusions. For example, analysis of the National Crime Victimization Survey [5] found few differences between weighted and unweighted regression coefficients, leading the authors to conclude that the weights were unnecessary in their case [6]. However, analysis of the National Maternal and Infant Health Survey [7] found the opposite, and the authors called for a consensus on the use of weights in regression analysis [8]. Over twenty years later, researchers using the National Health and Nutrition Examination Survey (NHANES) 2003-2004 [9] found little difference between weighted and unweighted regression coefficients, and chose to report the unweighted results [10].

The Canadian Longitudinal Study on Aging (CLSA) [11] is an ongoing complex survey of over 50,000 persons aged between 45 and 85 years. The CLSA collects clinical, social, psychological, lifestyle, economic, and genetic data from participants. Similar to NHANES [9] or the earlier Canadian Study of Health and Aging [12], the CLSA generates a large volume of secondary data analyses, and researchers who wish to make population-level inferences from the data will need guidance on the use of sample weights. Therefore, we examined the impact of the CLSA’s sample weights on a cross-sectional investigation of the association between social support availability (SSA) and two domains of cognitive function: memory and executive function. We chose this association because it is a component of our larger research program to use CLSA data and examine links between factors of social engagement, vulnerability, and cognitive function.

## Materials and Methods

### Study and Sample

The CLSA contains two components, known as the Comprehensive (n = 30,097) and Tracking (n = 21,241) Cohorts [13]. Baseline Comprehensive data were collected at in-home interviews and data collection site visits; baseline Tracking data were collected via telephone interviews. For both components, individuals were randomly selected in pre-specified provincial, age, and sex strata through random-digit dialling and mailouts from provincial health ministries [1]. Some Tracking participants were recruited from the pool of persons who completed the Canadian Community Health Survey on Healthy Aging [14]. Comprehensive participants were sampled from defined areas surrounding 11 data collection sites located in seven Canadian provinces; these defined areas had a radius of 25-50 kilometers around each site. Tracking participants had to reside in one of Canada’s ten provinces.

### Study Variables

We utilized baseline data from the Comprehensive component in our analyses to take advantage of the larger sample size compared to the Tracking component. Our analysis included five measures of cognitive function spanning the two cognitive domains: 1) Memory: Rey Auditory Verbal Learning Test Trial 1 and Rey 5-minute delayed recall [15]; and 2) Executive function: Mental Alternation Test [16], Animal Fluency Test [17], and Controlled Oral Word Association Test [18]. Details of the tests and scoring are available elsewhere [19,20].

We obtained z-scores (mean = 0; standard deviation = 1) for each cognitive test by subtracting the mean test score from participants’ raw scores and dividing the difference by the standard deviation of the mean test score. The means, standard deviations, and z-scores were computed separately for English and French speakers to adjust for language-based differences in test performance [20]. For each cognitive domain, we added the z-scores for the domain-specific tests together to obtain overall domain scores. For the Animal Fluency Test, we awarded one point for each distinct animal named, even if the animals were part of the same species group (e.g., Maltese and German shepherd would each receive one point, despite both being canines) [19].

We assessed SSA with the Medical Outcomes Study–Social Support Survey [21]. The survey asked participants to rate their level of perceived support in each of 19 different areas on a 5-point scale ranging from 1 (none of the time) to 5 (all of the time). The average score across these 19 areas formed an overall SSA index with a maximum score of 5 [22].

Covariates included sex, age group (45-54 years, 55-64 years, 65-74 years, ≥ 75 years), province of residence, education (completed high school – yes/no), cigarette smoking (smoked at least 100 cigarettes over a lifetime– yes/no), alcohol consumption in the past 12 months (at least weekly, 2-3 times monthly or less, never), self-reported hypertension (yes/no), self-reported diabetes/borderline diabetes/high blood sugar (yes/no), depressive symptoms (measured on a 0 [low] to 30 [high] scale with the 10-item Center for Epidemiologic Studies Depression Scale [23]), any level of help required on at least one of seven basic activities of daily living (ADL) versus no help required, and any level of help required on at least one of seven instrumental activities of daily living (IADL) versus no help required. The covariate set was chosen in accordance with the CLSA’s recommendation to include sex, age group, and province of residence in regression models, as well as with the findings of previous research into social support and cognition-related outcomes [1,24,25]. ADLs and IADLs were assessed using the Older Americans Resources and Services Multidimensional Functional Assessment Questionnaire [26].

### Unweighted and Weighted Regression Analyses

We used multiple linear regression to examine the association between SSA and each cognitive domain separately, controlling for the covariates mentioned above. We also tested for an interaction between SSA and age group because of the likelihood of a strong relationship between age and cognitive function. Within each cognitive domain, we ran two regression models, one unweighted and the other weighted.

We used PROC GENMOD in SAS v9.4 (SAS Institute, Cary, NC) to conduct the unweighted regressions and PROC SURVEYREG in SAS to conduct the weighted regressions. PROC SURVEYREG can handle survey weights to address the non-representativeness of the sample, while other procedures have weight statements to correct for heterogeneous residuals. In the programming syntax for PROC SURVEYREG, we specified the CLSA’s weight variable (WGHTS_ANALYTIC_COM) in the weight statement, the geographic strata variable (WGHTS_GEOSTRAT_COM) in the strata statement, and the unique identification variable for participants in the cluster statement [1]. The analytic weights were constructed from the inflation weights to have a mean weight of one within each stratum and thus add up to the CLSA sample size, while the inflation weights, which were developed from the design probabilities of including participants, add up to the complete population size.

The primary regression analyses excluded observations with missing data. We also conducted multiple imputation to examine the effect of missing data on the regression results. The issue of missing data was particularly important because the weights were calculated using the entire sample, whereas only complete cases were included in the primary regression models. The prediction variables for the imputation procedure included all the variables used in the regression models described above.Fully conditional specification methods—linear regression for continuous variables and logistic regression for categorical variables—involving 20 burn-in iterations generated five imputed datasets that were individually re-analyzed using the regression procedures described above. The regression results were pooled across the imputed datasets to yield single sets of regression coefficients and standard errors for each model.

### Simulation Study

The simulation study mimicked CLSA data in four steps. In the first step, we calculated new weights by multiplying the CLSA analytical weights with a lognormally distributed random variable. This distribution was chosen to ensure all weights were positive with a median value of 1 (not changing the sample size).

Instead of using these simulated weights in the regression analysis later, we used them to generate ‘true’ regression coefficients (reverse engineering). In this way, we could keep the CLSA analytical weights fixed and view them as estimates from these true simulated weights. In the second step, we used weighted regression (PROC SURVEYREG) with the simulated weights on the original CLSA data to generate the true coefficients (*β*_*TRUE*_) of all the variables included in the regression models described above. In the third step, we simulated covariates and exposures using a normal copula (a probability model representing the joint probabilities of a multivariate distribution) that was estimated from the original CLSA data. Through this process, we simulated new participants for CLSA who were like those observed in CLSA. We did not simulate new values for age, sex, education, and province because these variables were used to create the analytical weights that we fixed in the simulation. In the final step, we simulated data on the response variable using the regression model with the true regression coefficients from the first step and added residual noise.

The simulated CLSA data were then analyzed with and without the analytical weights. We examined the differences (bias) between the regression coefficients obtained from these simulated unweighted and weighted analyses with *β*_*TRUE*_. Using 1,000 simulations, we also estimated the proportion of the simulated regression coefficients whose 95% confidence intervals contained *β*_*TRUE*_ (coverage).

## Results

The sample size was 30,097 (51% female) with a median age of 62 years (range: 45 to 86 years). Further sample characteristics are shown in Table 1. The overall SSA index was left-skewed (median = 4.42; interquartile range = 0.95) (Fig 1). For the cognitive domains, the z-scores exhibited normality (Fig 2).

**Table 1.**
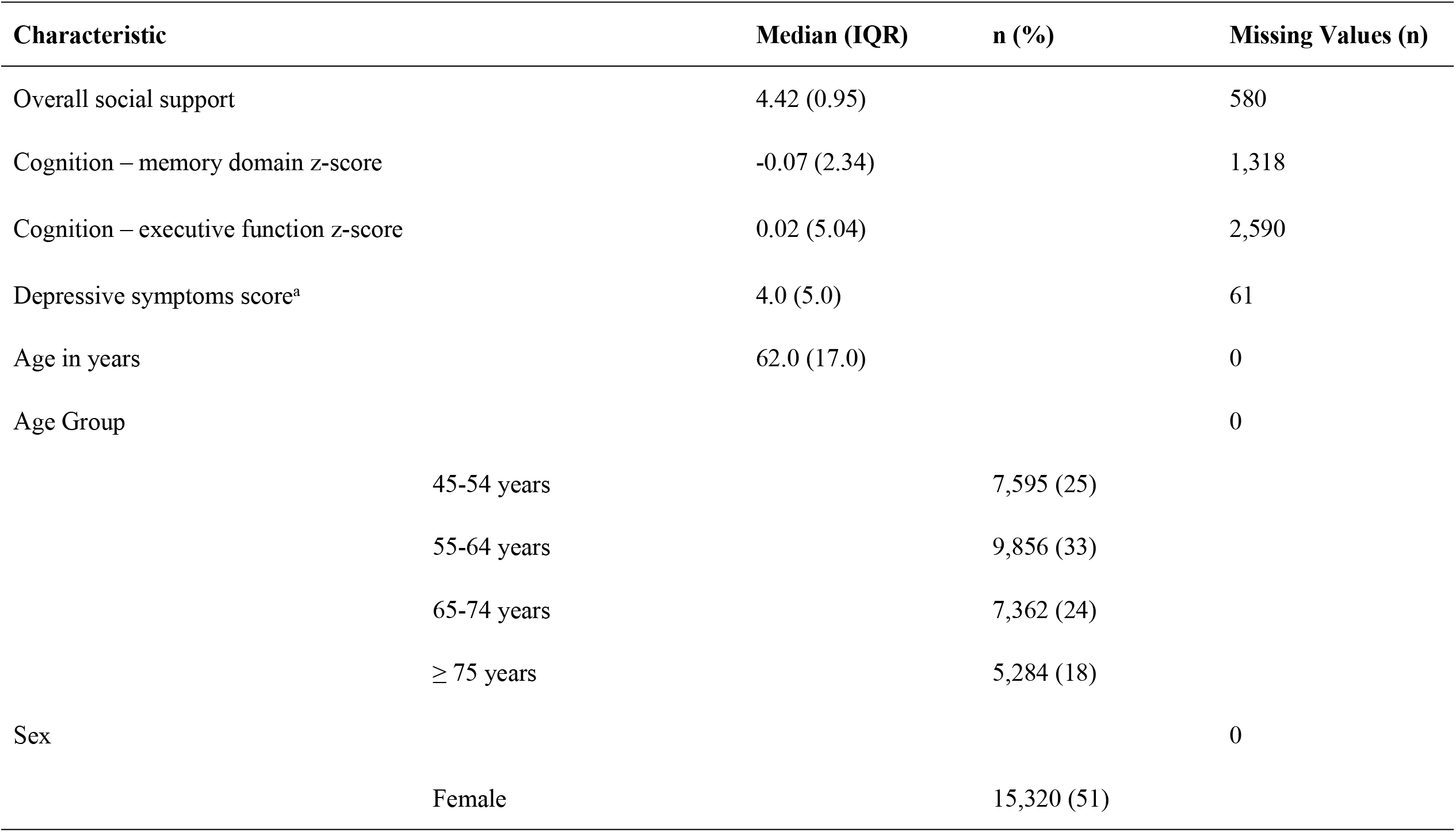
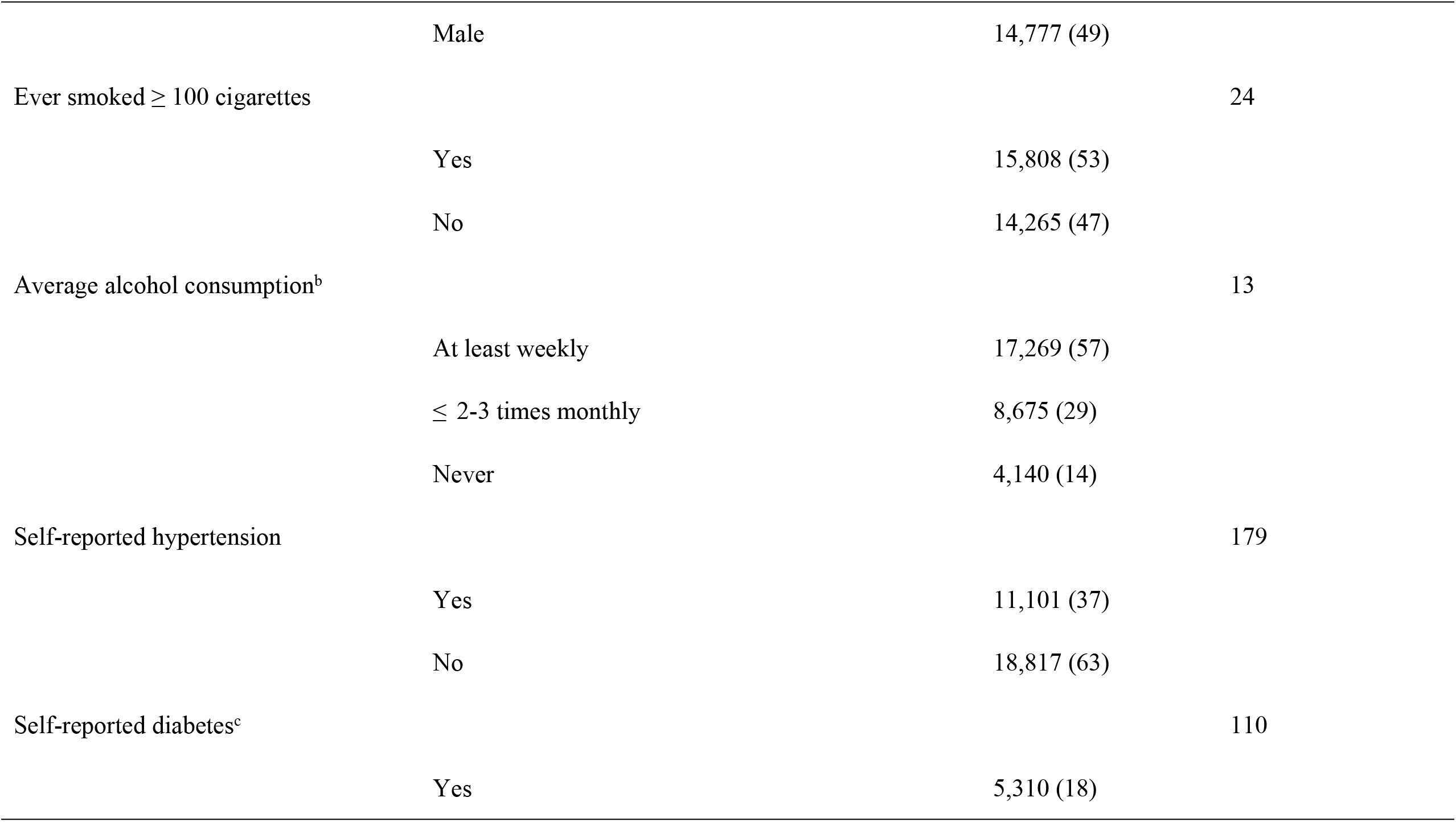
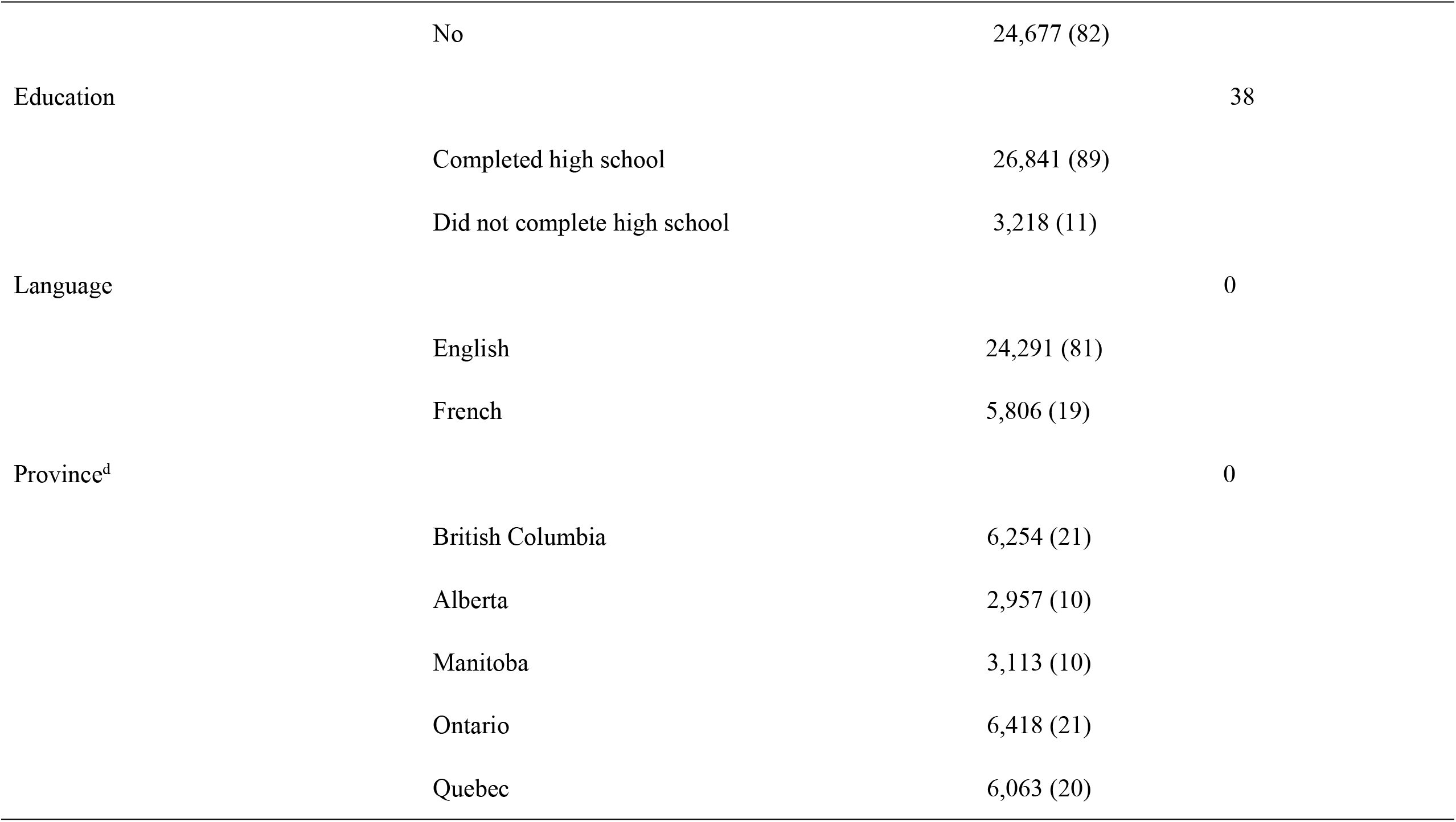
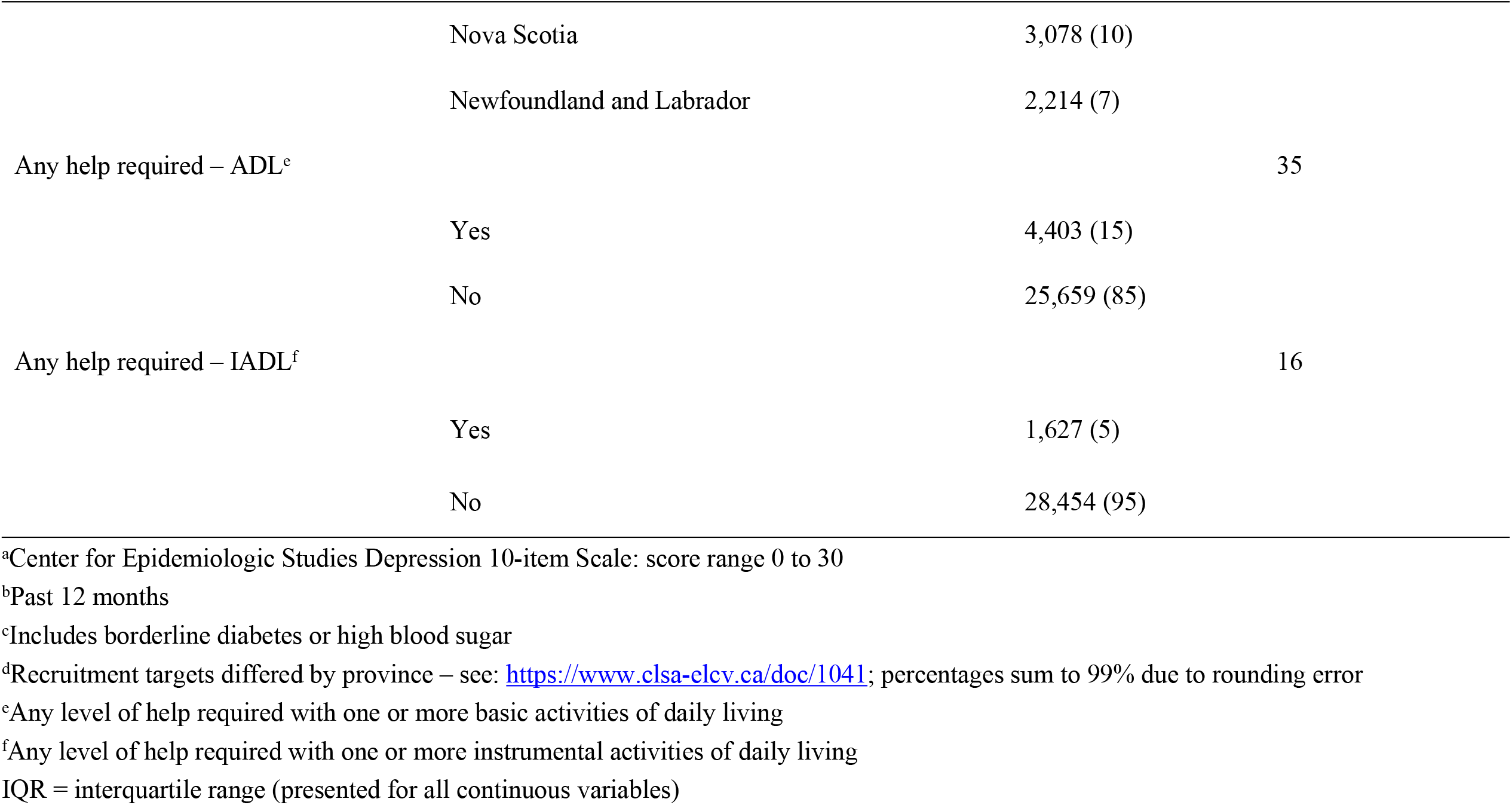
Sample Characteristics (n = 30,097).

**Fig 1.**
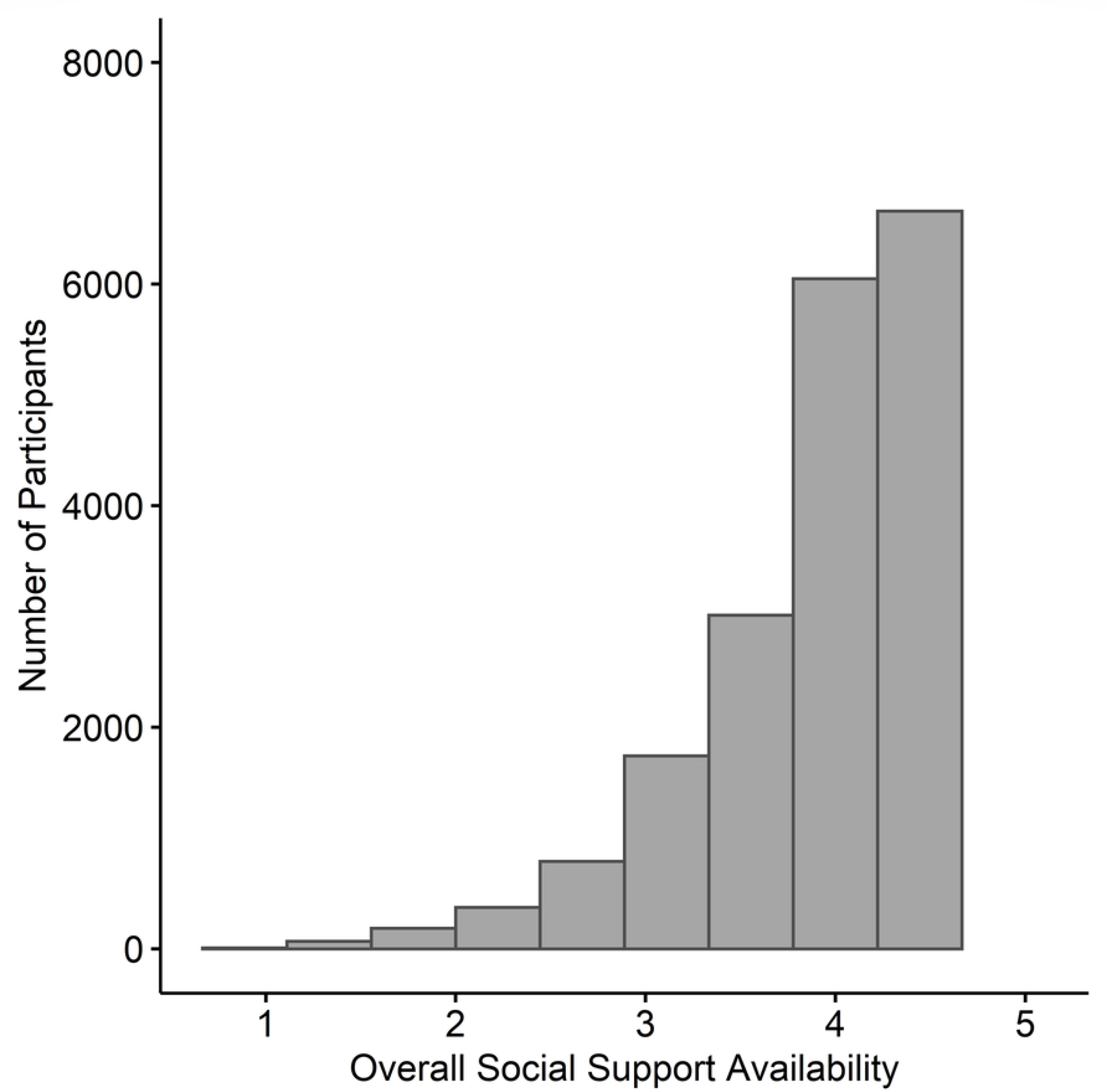
Distribution of Overall Social Support Availability Index. Distribution of overall social support availability. This score was calculated by averaging the scores for 19 different questions on the Medical Outcomes Study–Social Support Survey. For example, an overall index score of 4 indicates that a participant’s average score was 4 on the 19 questions. Higher scores indicate greater social support availability.

**Fig 2.**
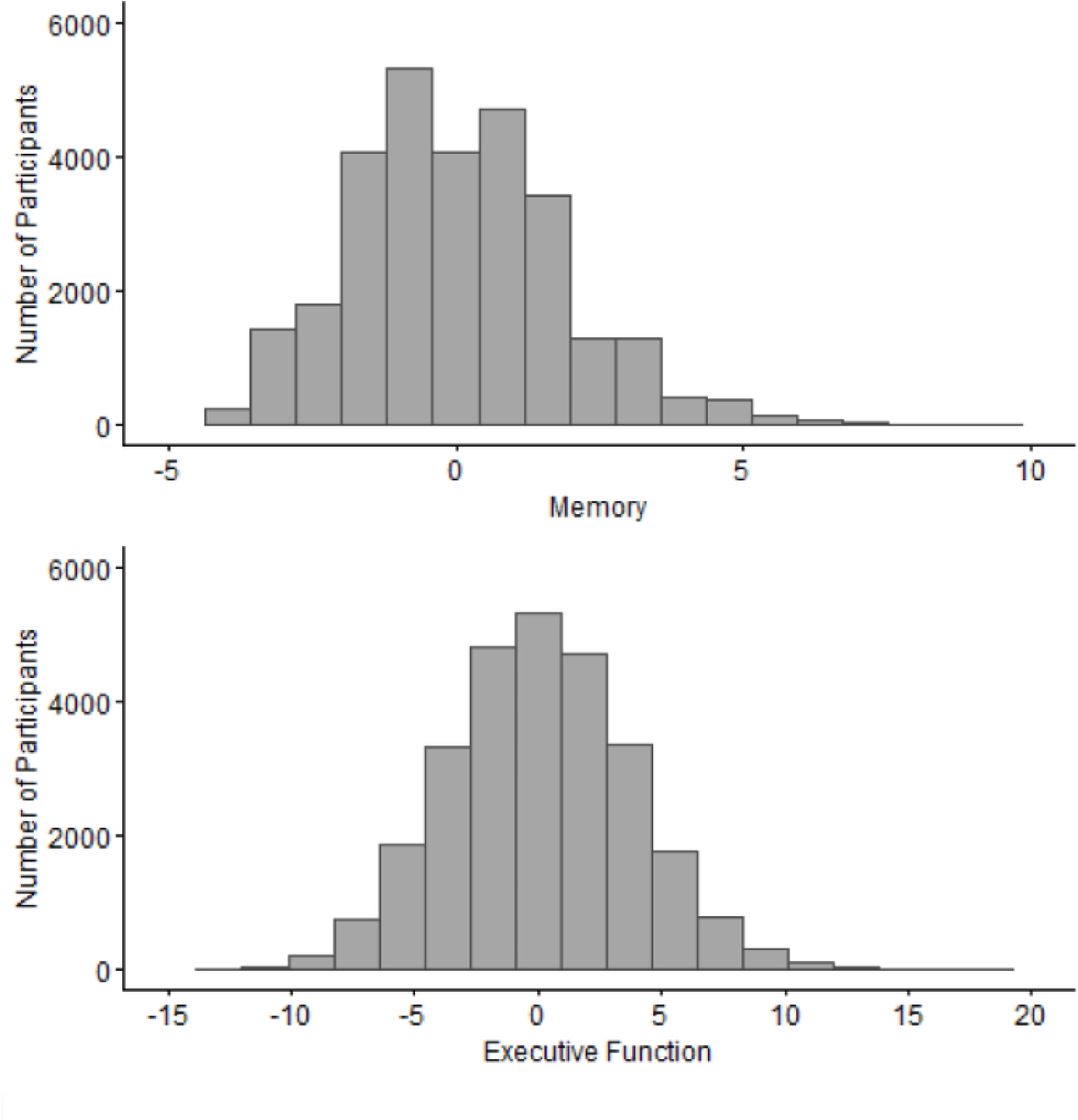
Distribution of Cognitive Function z-scores by Domain.

### Unweighted and Weighted Regression Analyses

All regression analyses showed a positive association between SSA and both domains of cognitive function (Table 2). Within each domain, the regression coefficients for the unweighted and weighted analyses were almost identical (memory: 0.1600 unweighted versus 0.1548 weighted [Δ = 0.0052]; executive function: 0.3607 unweighted versus 0.3613 weighted [Δ = -0.0006]). The standard errors across both domains were slightly smaller for the unweighted analyses (memory: 0.0154 unweighted versus 0.0181 weighted [Δ = -0.0027]; executive function: 0.0332 unweighted versus 0.0384 weighted [Δ = -0.0052]) (Table 2). None of the interaction terms between SSA and age group were statistically significant at the 5% level.

**Table 2.**
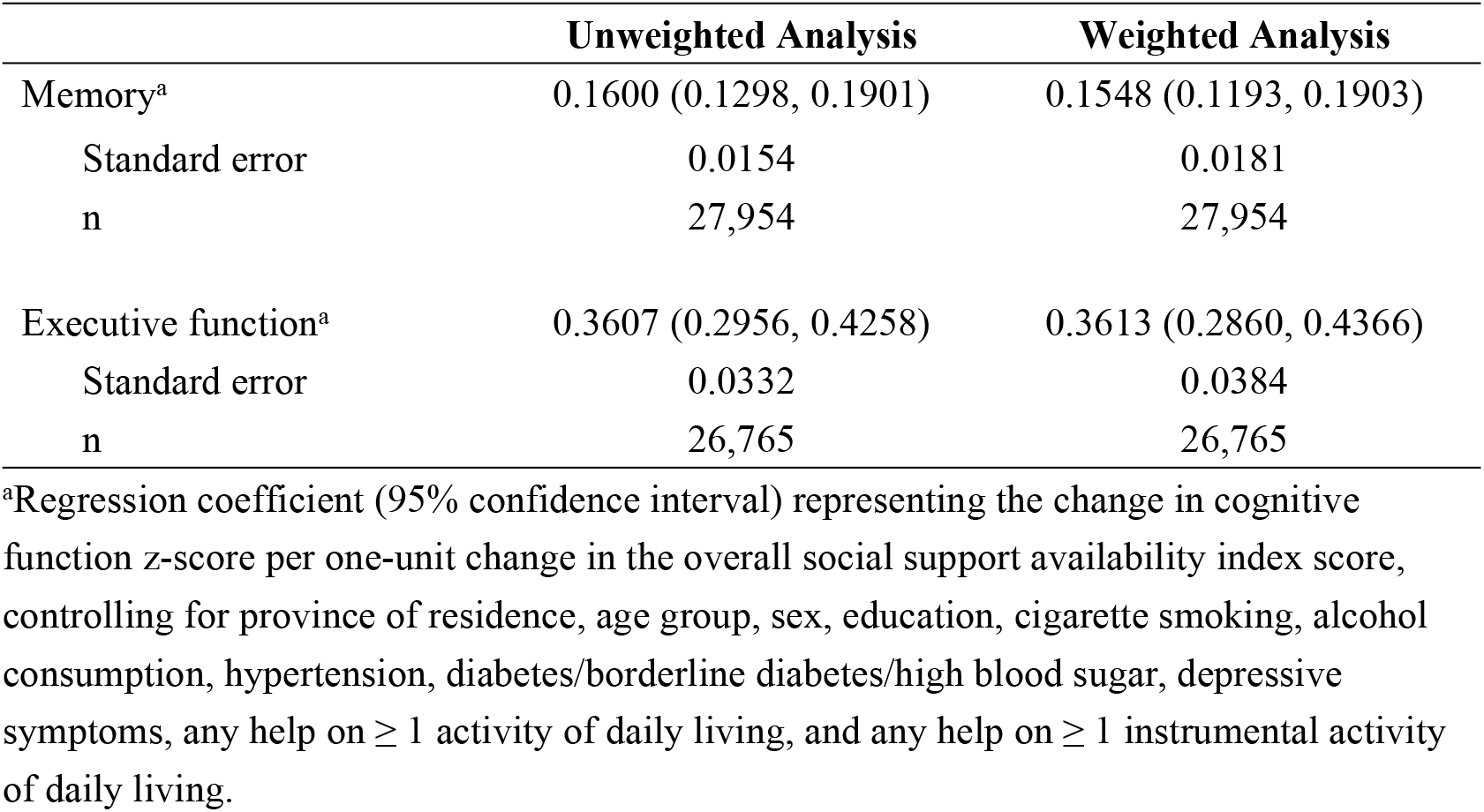
Unweighted and Weighted Multiple Regression Analyses.

Following multiple imputation, the regression coefficients and standard errors did not change appreciably compared to the complete case analyses.Differences in model parameters ranged from 0.0% to 3.3% (S1 Appendix).

### Simulation Results

The simulation results confirmed the existence of very little bias in regression coefficients when using either unweighted analysis (0.69 × 10^−4^ to 8.54 × 10^−4^) or weighted analysis (0.04 × 10^−4^ to 11.52 × 10^−4^), compared to *β*_*TRUE*_. In most cases, the weighted analysis had a smaller bias than the unweighted analysis, but both were minimal (Table 3). However, the standard errors from the unweighted analyses were about 10% less than those from the weighted analyses. The coverage of both the weighted and unweighted analyses was very close to 95%. Therefore, in approximately 95% of cases, *β*_*TRUE*_ fell within the confidence intervals of the simulated weighted and unweighted regression coefficients.

**Table 3.**
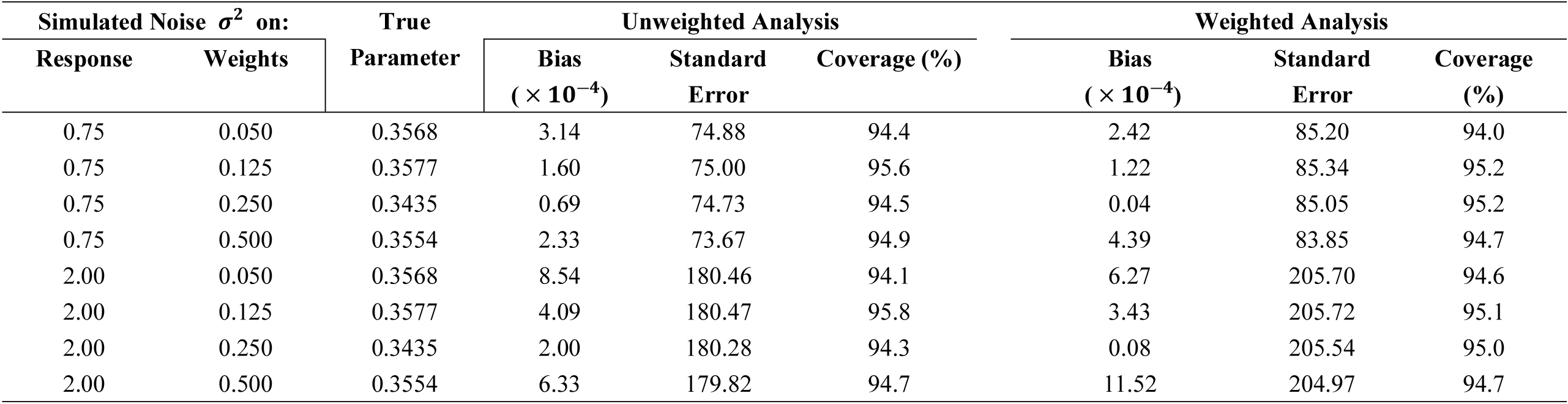
Bias, Standard Error, and Coverage Probability of the Exposure Variable in the Regression Analysis from the Simulation Study.

## Conclusions

Our results do not support requiring the use of weights when analyzing the association between SSA and cognitive function. This is because the unweighted and weighted analyses produced regression results that were similar to one another, despite the fact the unweighted analyses did not account for the complex survey sampling design of the CLSA. The simulation study confirmed these results and showed the standard errors of both analytic methods were appropriate (coverages for 95% intervals were nominal) and we did not observe a systematic underestimation of the population variance, as was seen in other work [4]. Since the unweighted analyses had smaller standard errors, we recommend the unweighted approach for analyses undertaken with CLSA data on SSA and cognitive function.

Our finding may not hold true for all associations between variables collected in the CLSA. Therefore, we recommend always conducting a sensitivity analysis using the sample weights and an appropriate statistical procedure (e.g., PROC SURVEYREG), and comparing the result to the unweighted analysis of the same data. When the results are similar, the unweighted analysis can be used as the primary analysis because the lower standard errors will result in smaller confidence intervals with adequate coverage. Conversely, when the results differ, we suggest researchers employ the weights in their primary analysis. Determining how different is ‘too different’ is a qualitative decision that may vary by research area. Thus, we suggest that the secondary analysis be reported as a sensitivity analysis in an appendix for transparency.

The large sample sizes underlying the results in Tables 2 and 3, as well as the CLSA’s effort to cover a large geographical portion of Canada, meant the CLSA’s participants represented the Canadian population. This may help explain why both analytical approaches provided similar results.

In our work, changes in variance when shifting from a weighted to an unweighted analysis affected the width of the confidence intervals, but did not affect the coverage of the confidence intervals. This meant both analytical approaches provided appropriate standard errors for the regression coefficients included in our models. Therefore, we were less concerned with incorrectly calling a finding ‘statistically significant’ in the unweighted analysis, which has been an issue in studies with smaller sample sizes [27].

Regarding limitations, we did not explore differences between unweighted and weighted analyses for all of the cognitive tests in the Comprehensive Cohort, nor did we analyze any data from the Tracking Cohort. Additionally, our work utilized baseline data and the findings might change for longitudinal analyses.

In conclusion, the current study shows that employing analytical weights would not be necessary for at least some association studies using CLSA data, although we cannot guarantee that this would be the case for all analyses of associations in CLSA.

## Data Availability

Researchers interested in accessing data from the Canadian Longitudinal Study on Aging must meet the criteria for access and submit a data access application. Further details are available at https://www.clsa-elcv.ca/access

## Acknowledgements

This research was made possible using the data collected by the CLSA. Funding for the CLSA was provided by the Government of Canada through the Canadian Institutes of Health Research under grant reference number LSA 94473 and the Canada Foundation for Innovation, as well as the following provinces, Newfoundland and Labrador, Nova Scotia, Quebec, Ontario, Manitoba, Alberta, and British Columbia. This research has been conducted using the CLSA dataset Comprehensive data v.3.6 (Baseline plus Cognition) under Application ID #1909009.

The authors thank Mary Thompson for her helpful comments on earlier drafts of this manuscript and Levi Prikken for conducting the simulation studies as part of his master’s thesis in mathematics at the Eindhoven University of Technology.

The opinions expressed in this manuscript are the authors’ own and do not reflect the views of the Canadian Longitudinal Study on Aging.

Data from the CLSA are not publicly available. However, researchers may submit a data access request to the CLSA and obtain data for their own analyses. Information on accessing CLSA data is available here: https://www.clsa-elcv.ca/data-access

## Supporting Information

**S1 Appendix. Multiple imputation versus complete case analysis**.

